# Exploring characteristics of visual search in older adults and people with Parkinson’s during adaptive gait

**DOI:** 10.64898/2026.05.12.26352982

**Authors:** Jiaxi Ye, Yuri Russo, Zijing Wang, Phaedra Leveridge, Sarah E Lamb, David Harris, Mark Wilson, William R. Young

## Abstract

Freezing of gait is a disabling episodic symptom of Parkinson’s disease, typically emerging during complex locomotor tasks such as turning, obstacle negotiation, and gait initiation. These tasks require effective motor planning and proactive visual search of the intended walking route. Current evidence suggests that people with Parkinson’s disease and freezing of gait show different patterns of visual search compared to those without freezing of gait and healthy older adults. However, existing reports are based on relatively simple tasks that lack common triggers for freezing of gait and do not adequately control for other factors likely to influence visual search, such as motor symptom severity and balance ability.

This study examined visual search behaviour in 24 healthy older adults and 37 people with Parkinson’s disease (21 with freezing of gait, 16 without) during a complex walking task requiring repeated turning and navigation through narrow spaces. Visual search characteristics were compared between people with Parkinson’s disease and healthy controls, and relationships between visual search, freezing of gait, motor symptom severity, and balance ability were explored within the Parkinson’s disease group.

Compared with healthy controls, people with Parkinson’s disease showed significantly fewer fixations toward areas outside the walking path, longer average fixation durations, and reduced saccade amplitudes, with no differences in proactive visual planning of the intended route. No relationship was found between visual search outcomes and freezing of gait. Reduced fixations to outside-path areas were associated with poorer functional balance independently of motor symptom severity.

These findings indicate that restricted visual sampling in Parkinson’s disease is primarily associated with balance impairment rather than freezing of gait or motor symptom severity.

## 1. Introduction

Visual information is critical for planning future stepping actions during adaptive locomotion (Matthis et al., 2018). In people with Parkinson’s disease (pwPD), visually complex constraints such as narrow doorways, cluttered spaces, and obstacles can disrupt locomotion and trigger freezing of gait (FOG), potentially through increasing visuospatial demands and perceptual load (Almeida and Lebold, 2010, Beck et al., 2015). In contrast, structured visual cues can improve gait performance and reduce FOG by providing spatial references that support movement planning (Ginis et al., 2018). These findings highlight the functional relevance of visual information for adaptive walking in people with PD and underscore the importance of understanding how visual search behaviour is altered in this population.

A growing body of work indicates that eye movement behaviour is systematically altered in pwPD, represented by longer fixation durations and reduced saccade amplitudes (Gibbs et al., 2024; Matsumoto et al., 2011; Tsitsi et al., 2023). These oculomotor changes suggest a narrower spatial sampling strategy and slower updating of visual information, which may constrain the proactive visual search required to negotiate complex environments safely. Evidence from naturalistic walking paradigms indicates that pwPD, particularly those with FOG, preferentially direct gaze toward proximal regions of the walking path and show reduced allocation of visual attention to distal locations (Hardeman et al., 2020; Vanegas-Arroyave et al., 2022). This shift toward visual sampling of proximal areas may limit the ability to preview upcoming environmental constraints, potentially compromising gait adjustments required for safe locomotion.

Similar restricted proactive visual search has also been observed in older adults who experience fear of falling. Increased fall-related anxiety is associated with prioritisation of immediate stepping constraints at the expense of previewing future path information required for adaptive gait (Ellmers, Cocks, & Young, 2020; Ellmers et al., 2021). This convergence of findings suggests that reduced proactive sampling of the intended walking route may reflect an adaptive response to perceived instability or increased task demands. In pwPD, impaired balance and reduced automaticity of motor control are prevalent, particularly in individuals with FOG (Bryant et al., 2014; Lord et al., 2020; Wu et al., 2015). It is therefore plausible that altered visual search behaviour reflects an interaction between disease-related motor impairment and functional balance capacity, rather than FOG pathology alone.

Despite increasing interest in visual control of gait in PD, proactive visual search to distal locations (i.e., avoidance of fixating areas more than ∼2 steps ahead) has largely been examined during relatively simple locomotor tasks, such as straight-line unconstrained walking (Vanegas-Arroyave et al., 2022). These paradigms cannot represent visual sampling strategies during more complex adaptive walking conditions that require continuous processing of multiple environmental features. Furthermore, previous comparisons between pwPD with and without FOG have often relied on between-group contrasts without explicitly accounting for variation in overall motor severity, limiting interpretation of whether observed differences are specific to FOG pathology or reflect broader disease-related impairment (Russo et al., 2022; Vanegas-Arroyave et al., 2022).

In addition to proactive visual sampling of the walking path, fixation behaviour directed toward task-irrelevant locations (“outside area” fixations) may provide insight into cognitive load during locomotion. Experimental evidence indicates that healthy adults transiently disengage gaze from the walking path when attentional resources are allocated to concurrent mental arithmetic task (Ellmers et al., 2016; Russo et al., 2025). Such fixations may therefore reflect available cognitive capacity or flexibility in visuomotor control. Given the increased attentional demands associated with walking in PD (Kelly et al., 2012; Pieruccini-Faria et al., 2014; Raffegeau et al., 2019), reduced frequency of fixations outside the immediate walking path may indicate reduced spare cognitive capacity or increased prioritisation of gait stability. To date, this behaviour has not been systematically examined during complex adaptive walking in pwPD.

The first objective of this study was to determine whether pwPD exhibit altered visual search behaviour during complex adaptive walking compared with healthy older adults, with a specific focus on proactive visual search along the walking path and the occurrence of outside area fixations. Our secondary and primary objective was to determine whether individual differences in visual search behaviour within the PD group were associated with clinically relevant functional characteristics, including FOG severity (NFOG-Q, (Nieuwboer et al., 2009)), motor symptom severity (MDS-UPDRS-III, (Goetz et al., 2008)), and balance ability (Mini-BESTest (Franchignoni et al., 2018)). We hypothesised that pwPD would demonstrate reduced proactive visual search and fewer outside fixations compared with healthy controls. Within the PD group, we further hypothesised that reduced proactive visual search would be associated with poorer balance ability and greater FOG severity, independent of overall motor symptom severity.

## 2. Methodology

### 2.1. Participants

Sixty-one participants were included: 37 people with Parkinson’s disease (21 with FOG, 16 without FOG) and 24 age-matched healthy controls (HC).Participants were recruited through UK Parkinson’s support groups and community networks.

Sample size was determined a priori to detect between-group differences in visual search behaviour between pwPD and HC. Previous work has reported significant differences in visual search during walking in pwPD but did not report effect sizes (Vanegas-Arroyave et al., 2022). Based on consistently observed alterations in oculomotor behaviour in PD, including longer fixation duration and reduced saccade amplitude (Gibbs et al., 2024), a moderate-to-large effect size (Cohen’s d = 0.65) was assumed. Power analysis (α = 0.05, 1–β = 0.80) indicated a required sample of 21 participants per group.

Exclusion criteria were: (1) cognitive impairment (Montreal Cognitive Assessment (MoCA) score < 21/30) (Nasreddine et al., 2005), and (2) any neurological, musculoskeletal, or vestibular disorder other than PD affecting gait or balance. Ethical approval was obtained from the University of Exeter Department of Public Health and Sport Sciences Ethics Committee (21-12-08-B-02). All participants provided written informed consent prior to participation.

### 2.2. Clinical and cognitive assessment

Motor symptom severity in pwPD was assessed using Part III of the Movement Disorder Society Unified Parkinson’s Disease Rating Scale (MDS-UPDRS-III), with higher scores indicating greater motor impairment (Goetz et al., 2008).

Freezing of gait was assessed using the New Freezing of Gait Questionnaire (NFOG-Q) (Nieuwboer et al., 2009). Presence of FOG was determined using Item 1 of the NFOG-Q (self-reported occurrence of freezing episodes). FOG severity was quantified using the total NFOG-Q score, with higher scores indicating more severe and frequent freezing.

Dynamic balance was assessed in both groups using the Mini-Balance Evaluation Systems Test (Mini-BESTest), where higher scores indicate better balance performance (Franchignoni et al., 2018).

Attentional and psychological factors were assessed using the Gait-Specific Attentional Profile (G-SAP) (Young et al., 2020), which includes four subscales: Conscious Movement Processing (CMP), Anxiety/Arousal, Task-Irrelevant Worry (TI), and Processing Inefficiency (PI). Following recent psychometric validation in PD populations (Rosenblum et al., 2025), Item A2 (“concern about others’ evaluation”) was removed from the Anxiety subscale to improve construct validity; this modified subscale is referred to as G-SAP Arousal. General anxiety and depression symptoms were assessed using the Hospital Anxiety and Depression Scale (HADS), comprising anxiety (HADS-A) and depression (HADS-D) subscales (Zigmond & Snaith, 1983).

### 2.3. Experiment Protocols

Participants with Parkinson’s disease (pwPD) performed the walking task while optimally medicated (“ON” state). Medication retake was permitted where required to ensure testing reflected typical daily functioning. The task was conducted on a 3.6 m × 3.6 m platform (Figure 1). Circular yellow–black striped turn targets (TTs) were positioned at the midpoint of each side of the platform. Upon stepping on each TT, participants performed two 360° turns in alternating directions (a modified version of the protocol described as Ziegler test here (Goh et al., 2022)), with initial turning direction self-selected. Inner (yellow) and outer (blue) cones were positioned to form a narrow pathway between successive TTs. Inner cones were aligned with the upcoming TT and represented proximal obstacles requiring foot placement. Participants completed two full laps of the platform in both clockwise and anticlockwise directions. If needed, rest breaks were permitted at any time.

**Figure 1:**
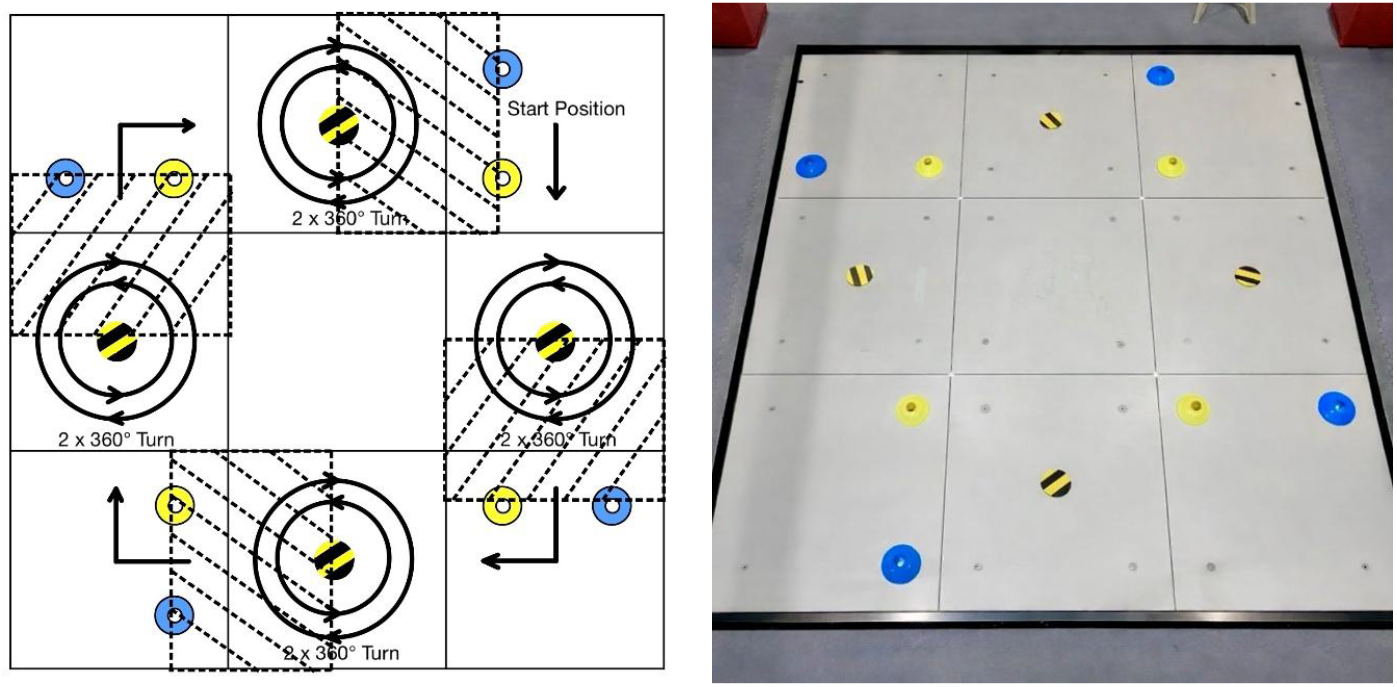
Protocol of the walking task. Arrows indicate the walking and turning directions. Inner (yellow) and outer (blue) cones are placed to create a narrow passage. The black and yellow striped targets represent the location where participants were instructed to stop and complete the turning tasks.

### 2.4. Equipment and metrics

Eye movements were recorded using Tobii Pro Glasses 2 (Tobii, Stockholm, Sweden). Binocular gaze data were sampled at 100 Hz and scene video was recorded at 25 Hz with a resolution of 1920 × 1080 pixels. Eye-tracking data were analysed using Tobii Pro Lab (version 1.145.28180). Areas of interest (AOIs) were manually defined frame-by-frame within the scene video. Fixations and saccades were identified using the Tobii Pro Lab Velocity-Threshold Identification (I-VT) filter with a velocity threshold of 30°/s and minimum fixation duration of 60 ms. Walking behaviour was additionally recorded using a Sony HDR-CX405 digital camcorder (Sony Corporation, Tokyo, Japan) at 25 Hz to support behavioural verification of task events.

Eye-movement outcomes were calculated within predefined time windows corresponding to visually guided walking segments (Figure 1). The time window of interest was defined as the interval between completion of each 360° turn and subsequent passage between the cone obstacles, representing periods requiring proactive visual guidance of gait.

Five AOIs were defined as follows: the turn target (TT), indicative of proactive visual search for upcoming locomotor goals; the platform, representing the walking surface; the inner obstacle, a proximal cone (yellow) aligned with the upcoming TT; the outer obstacle, a lateral boundary cone (blue); and the outside area, comprising regions beyond the walking platform that were considered to be task-irrelevant.

For each AOI, five eye-movement metrics were computed to capture complementary components of visual search behaviour, including the spatial allocation of attention and gaze transition dynamics (Cullipher et al., 2018). Number of fixations per second was derived by dividing the number of fixations within an AOI by the duration of the time window. Average fixation duration (ms) and total fixation duration (ms) indexed the average and cumulative dwell time per AOI, respectively. The percentage of total fixation duration expressed each AOI’s cumulative dwell time as a proportion of fixation duration summed across all AOIs. Finally, average saccade amplitude (degrees) quantified the mean angular displacement between successive fixations. Together, these measures provide established indices of visual sampling behaviour, reflecting the spatial distribution and temporal characteristics of attentional allocation during locomotion.

### 2.5. Statistical analysis

Statistical analyses were conducted using IBM SPSS Statistics (version 28; IBM Corp., Armonk, NY, USA). For the primary objective, between-group differences (pwPD vs HC) in visual search variables were assessed.Normality was evaluated using Shapiro–Wilk tests and homogeneity of variance using Levene’s tests. Welch’s t-tests were used for normally distributed variables, as this approach is robust to unequal variances. Mann– Whitney U tests were applied where normality assumptions were violated.For the secondary objective, associations within the pwPD group between visual search outcomes and clinical variables (FOG severity, motor symptom severity (MDS-UPDRS-III), and balance ability (Mini-BESTest)) were examined using Spearman correlation coefficients. Partial Spearman correlations controlling for MDS-UPDRS-III scores were conducted where appropriate to determine whether observed relationships were independent of overall motor severity.

Exploratory analyses examined associations between basic oculomotor characteristics and psychological measures, including anxiety (HADS-A) and conscious movement processing (G-SAP CMP), using Spearman correlations. Statistical significance was set at p = 0.05. To control for multiple comparisons, p-values were adjusted using the Benjamini–Hochberg false discovery rate (FDR) procedure (Benjamini & Hochberg, 1995). Effect sizes for between-group comparisons were reported as rank-biserial correlation (r), and correlation strength was expressed using Spearman’s rho (r).

## 3. Results

### 3.1. Demographic and clinical characteristics

A total of 67 participants were recruited and completed the experimental protocol: 25 HC, 19 pwPD without FOG, and 23 pwPD with FOG. Following data collection, six participants were excluded from the analysis: one HC and two pwPD without FOG and three pwPD with FOG were excluded owing to insufficient eye-tracking data quality (e.g., poor calibration or excessive signal loss). The final analysed sample therefore comprised 61 participants: 24 HC, 17 pwPD without FOG, and 20 pwPD with FOG (37 pwPD in total). Demographic and clinical characteristics of the analysed sample, stratified by group, are summarised in Table 1.

**Table 1:**
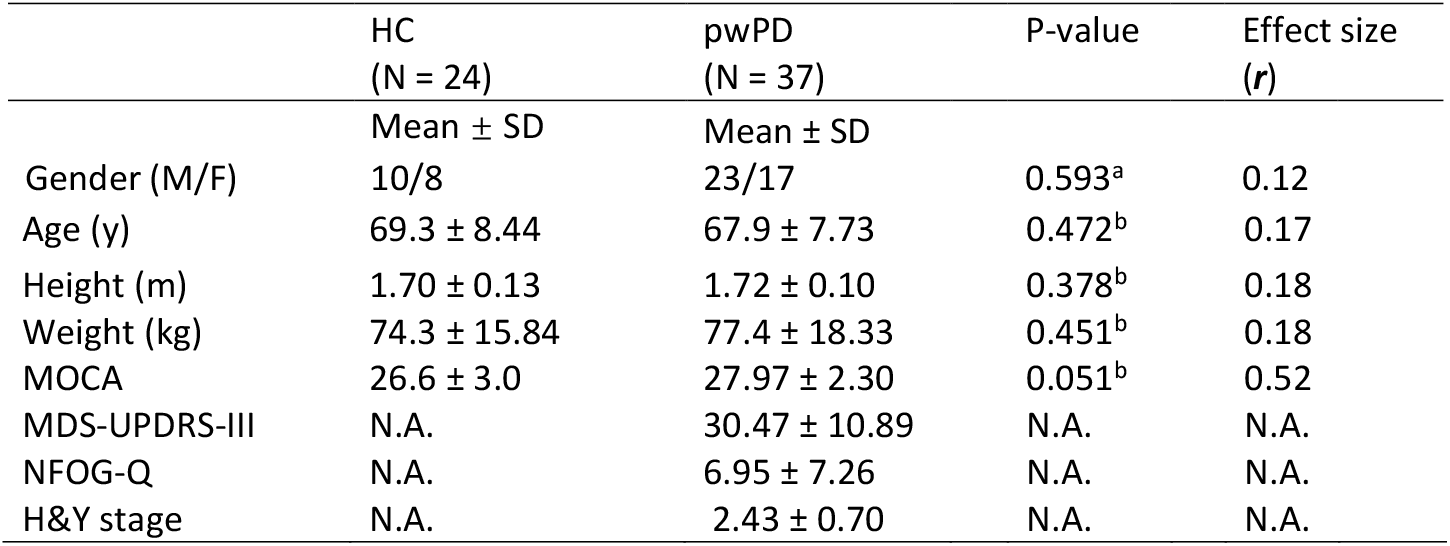
Demographics and clinical characteristics of health age-matched controls, people with PD. Significant differences (p< .05) are shown with *. (a) Gender distribution was assessed with Pearson’s Chi-square-test. (b) p-value of Student’s t-tests.

| | HC<br>(N = 24) | pwPD<br>(N = 37) | P-value | Effect size<br>($r$) |
| --- | --- | --- | --- | --- |
| | Mean ± SD | Mean ± SD | | |
| Gender (M/F) | 10/8 | 23/17 | 0.593<sup>a</sup> | 0.12 |
| Age (y) | 69.3 ± 8.44 | 67.9 ± 7.73 | 0.472<sup>b</sup> | 0.17 |
| Height (m) | 1.70 ± 0.13 | 1.72 ± 0.10 | 0.378<sup>b</sup> | 0.18 |
| Weight (kg) | 74.3 ± 15.84 | 77.4 ± 18.33 | 0.451<sup>b</sup> | 0.18 |
| MOCA | 26.6 ± 3.0 | 27.97 ± 2.30 | 0.051<sup>b</sup> | 0.52 |
| MDS-UPDRS-III | N.A. | 30.47 ± 10.89 | N.A. | N.A. |
| NFOG-Q | N.A. | 6.95 ± 7.26 | N.A. | N.A. |
| H&Y stage | N.A. | 2.43 ± 0.70 | N.A. | N.A. |

### 3.2. Group Comparison

#### 3.2.1. Proactive visual search and task-relevant obstacles/platform

No between-group differences were observed in the percentage of total fixation duration directed toward the turn targets (TT) (U = 404, p= .560, r = –0.09) or toward obstacle and platform regions (outer obstacle: U = 398, p = 0.492, r = –0.10; inner obstacle: U = 454.5, p = 0.881, *r* = 0.02; platform: U = 336.0, p = 0.112, *r* = –0.24). The number of fixations per second on the platform was also comparable between groups (t = 0.797, p = 0.428, d = 0.21).

#### 3.2.2. Fixations toward task-irrelevant locations (outside areas)

The pwPD group demonstrated fewer number of fixations per second directed toward outside areas compared with HC (U = 598, p = 0.023, r= 0.35). However, the percentage of total fixation duration allocated to the outside areas did not differ significantly between groups (outside areas: U = 552.0, p = 0.112, r = 0.24).

#### 3.2.3. Basic eye movements outcomes

Across all areas of interest, pwPD exhibited shorter average fixation durations than HC (U = 228.0, p < 0.001, *r* = –0.93). Saccade amplitude was also reduced in pwPD compared with HC (t = 2.302, p = 0.025, *d* = 0.60).

**Figure 2:**
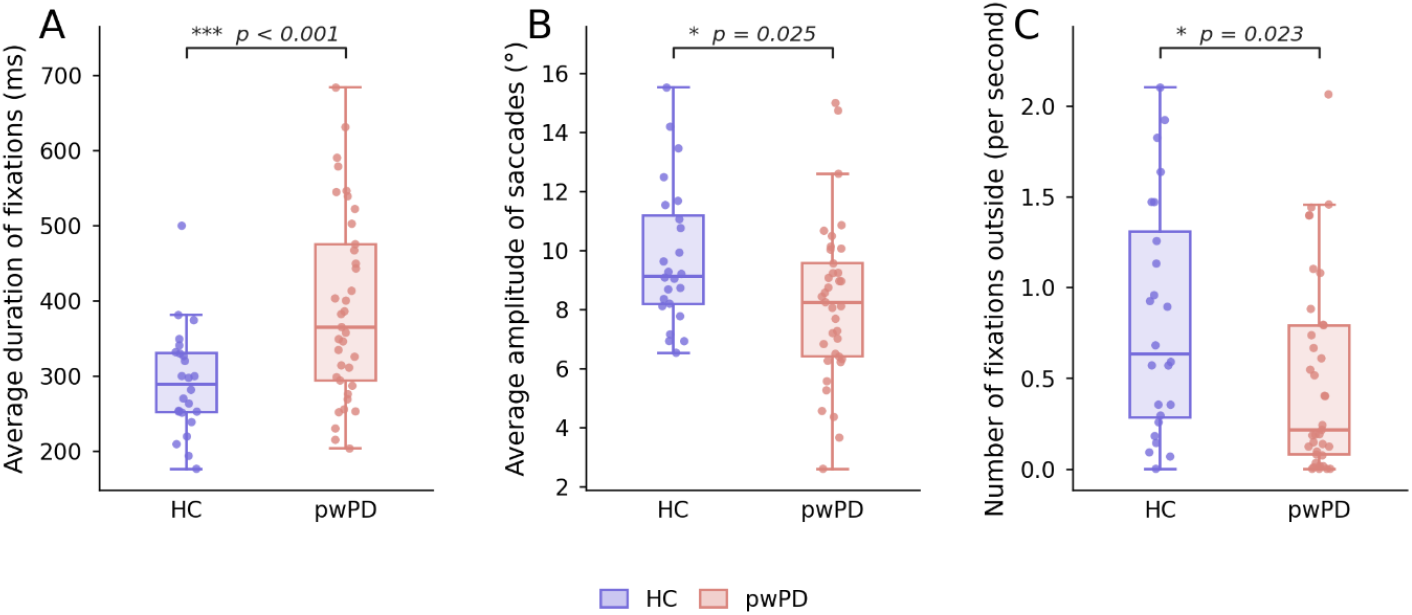
Group differences in oculomotor behaviour between healthy controls (HC) and people with Parkinson’s disease (pwPD). Box-and-whisker plots showing the distribution of (A) average fixation duration (ms), (B) average saccade amplitude (°), and (C) number of fixations outside area (per second) for healthy controls (HC; n= 24) and people with Parkinson’s disease (pwPD; n = 37). Boxes represent the interquartile range with the median indicated by the horizontal line; whiskers extend to 1.5 times the interquartile range. Individual data points are overlaid with horizontal jitter for clarity. Group comparisons were performed using an independent samples t-test (A, B) following confirmation of normality via the Shapiro–Wilk test, and a Mann–Whitney U test (C) where the assumption of normality was violated in the pwPD group. ^*^p < 0.05; ^***^p < 0.001.

### 3.3. Associations between visual search behaviour and clinical characteristics in pwPD

Within the pwPD group, functional balance ability (Mini-BESTest) was positively associated with visual disengagement from the walking path. Specifically, better balance performance was associated with a greater number of fixations per second directed toward outside areas (r = 0.512, p = 0.019) and a greater percentage of total fixation duration allocated to outside areas (r = 0.451, p = 0.033). No significant associations were observed between basic oculomotor characteristics (average fixation duration or average saccade amplitude) and FOG severity (NFOG-Q) or motor symptom severity (MDS-UPDRS-III).

After controlling for motor symptom severity (MDS-UPDRS-III), the associations between balance ability and outside area visual sampling remained statistically significant (number of fixations per second: r = 0.501, p = .029; percentage fixation duration: r = 0.450, p = 0.039). No other visual search variables showed significant associations with Mini-BESTest or NFOG-Q scores.

**Figure 3:**
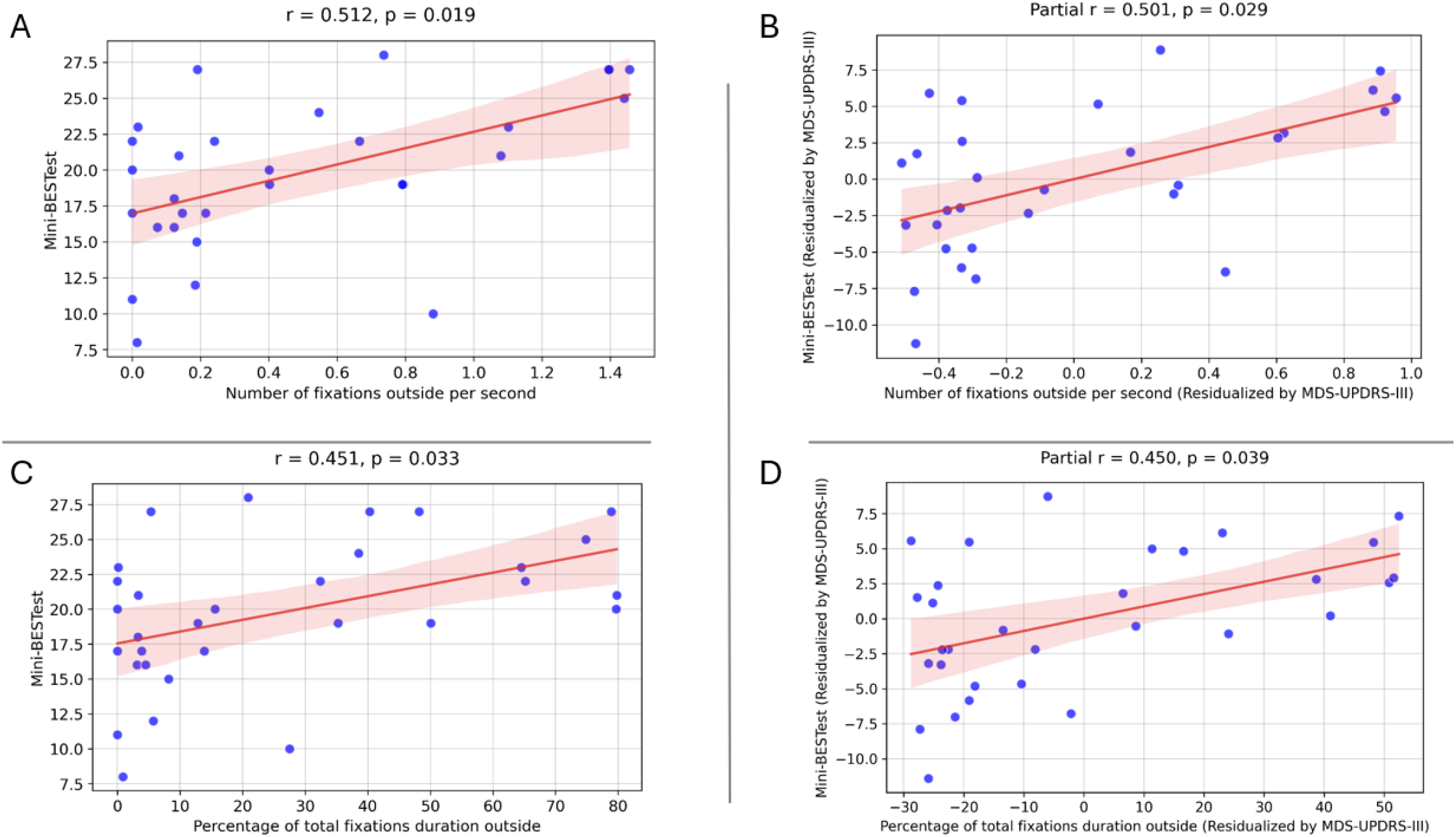
Correlations between visual search behaviour outside the target area and balance performance in people with Parkinson’s disease. Scatter plots illustrate the relationships between (A) the number of fixations outside the target area (per second) and Mini-Balance Evaluation Systems Test (Mini-BESTest) score, and (C) the percentage of total fixation duration outside the target area and Mini-BESTest score. Panels (B) and (D) display the corresponding partial correlations after controlling for disease severity (Movement Disorder Society-Unified Parkinson’s Disease Rating Scale Part III; MDS-UPDRS-III), using residualised scores for both the oculomotor and balance variables. Solid lines represent ordinary least-squares regression fits; shaded regions indicate 95% confidence intervals. All reported p-values are corrected for multiple comparisons using the Benjamini–Hochberg procedure. r, Pearson correlation coefficient; Partial r, partial correlation coefficient.

### 3.4. Exploratory associations with psychological measures

Group comparisons of G-SAP and HADS scores are reported in the supplementary materials. Within the pwPD group, reduced average saccade amplitude was associated with higher gait-specific arousal (G-SAP Arousal) scores (r = −0.535, p = 0.003). Higher depressive symptoms (HADS-D) were associated with longer average fixation duration (r = 0.387, p = 0.048). No significant associations were observed between fixation duration or saccade amplitude and other G-SAP subscales, including conscious movement processing (CMP) and processing inefficiency.

**Figure 4:**
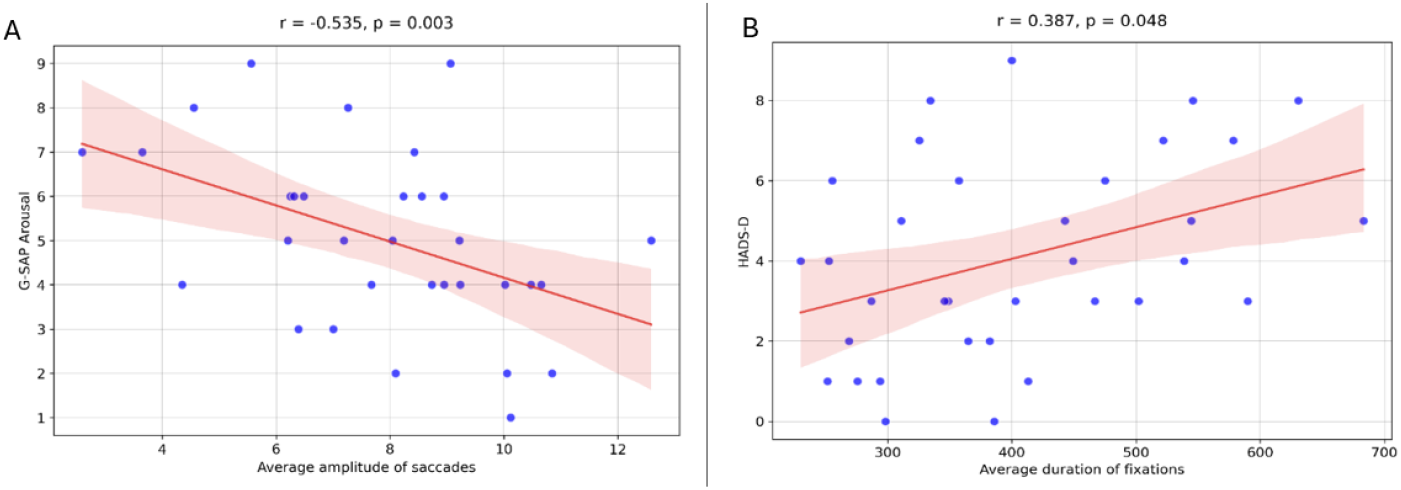
Associations between affective symptoms and oculomotor behaviour in people with Parkinson’s disease (pwPD). Panels (A): Relationship between G-SAP Arousal scores and average saccade amplitude (degrees) in pwPD. Higher gait-specific anxiety-related arousal was significantly associated with reduced saccade amplitude. Panels (B): Relationship between Hospital Anxiety and Depression Scale – Depression subscale (HADS-D) scores and average fixation duration (ms) in pwPD. Higher depressive symptom severity was significantly associated with longer fixation durations.

## 4. Discussion

The present study examined visual search behaviour during adaptive walking in pwPD and healthy controls. Contrary to our expectations, pwPD did not demonstrate reduced proactive visual search toward future stepping targets compared to HC. However, pwPD exhibited distinct oculomotor characteristics, including longer fixation durations, reduced saccade amplitudes, and fewer fixations directed toward task-irrelevant outside areas. Within the pwPD group, fewer outside fixations were associated with poorer balance performance but were not related to FOG severity or overall motor impairment. These findings suggest that narrowing of visual exploration during adaptive walking is more closely linked to functional balance capacity than to FOG pathology in pwPD.

### Proactive visual search toward turn targets

We observed no group difference in proactive fixation on the turn targets (TT). It is important to acknowledge that the TT location was predictable and required only modest stepping adjustments, meaning requirements for proactive acquisition of visual information regarding target position/orientations were perhaps reduced compared to other adaptive gait tasks used to evaluate visual search behaviour (Matthis et al., 2018) and could have reduced the sensitivity of outcomes to detect potential differences between groups (Takahashi et al., 2024).

### Reduced visual disengagement from the waking path

PwPD demonstrated fewer fixations toward task-irrelevant outside areas compared with HC. Importantly, the number of outside fixations was associated with poorer functional balance; an association that remained significant after controlling for motor symptom severity. In contrast, outside fixations were not associated with FOG severity. These findings indicate that reduced visual exploration beyond the walking path is more closely linked to balance capacity than to global disease progression or freezing pathology.

We argue that outside fixations broadly represent visuomotor flexibility during locomotion. Individuals with better functional balance may tolerate larger gaze shifts away from the immediate walking path without compromising postural stability (Russo et al., 2025). In contrast, individuals with reduced balance capacity may prioritise visual monitoring of task-irrelevant and proximal walking constraints in order to maintain stability (Young & Mark Williams, 2015). This interpretation is consistent with evidence that gaze shifts are mechanically coupled with postural control and can influence body sway dynamics (Kechabia et al., 2025). Larger amplitude gaze shifts may introduce instability, particularly in individuals with impaired sensorimotor integration. Reduced outside fixations may therefore reflect a conservative visual strategy that limits destabilising gaze shifts when postural control is challenged.

Importantly, the present findings do not demonstrate that outside fixations are necessarily functionally beneficial. Rather, they may emerge when task demands are sufficiently manageable to permit broader visual exploration. In this sense, outside fixations may reflect ‘spare’ visuomotor capacity rather than a strategy adopted to improve gait performance. Nevertheless, their association with balance ability suggests that outside fixation frequency may provide a relevant marker of functional stability during locomotion.

### Basic oculomotor alterations during adaptive walking

Consistent with previous laboratory-based studies, pwPD exhibited reduced saccade amplitude and longer fixation duration compared with HC. (Gibbs et al., 2024; Tsitsi et al., 2023; Yu et al., 2016)

Reduced saccade amplitude in PD is commonly attributed to altered basal ganglia–superior colliculus circuitry, resulting in increased inhibitory output that suppresses saccade generation (Antoniades & Spering, 2024; Pretegiani & Optican, 2017). Degeneration of dopaminergic neurons in the substantia nigra pars compacta disrupts the balance of direct, indirect, and hyperdirect basal ganglia pathways, leading to excessive inhibition of the superior colliculus and reduced saccade amplitude (Buhmann et al., 2015; Waldthaler et al., 2019). Such alterations may constrain the spatial extent of visual sampling, potentially limiting the acquisition of information required for gait adjustments.

Similarly, prolonged fixation durations may reflect slower disengagement of gaze or increased reliance on stable visual input when motor automaticity is reduced (Redgrave et al., 2010). Increased fixation duration has been interpreted as a strategy to maintain stable perceptual information when visuomotor control becomes less efficient (Ellmers, Cocks, Kal, et al., 2020). Together, these changes indicate a more restricted visual sampling strategy that may reduce flexibility in adapting gait to environmental constraints.

Importantly, these oculomotor characteristics were observed during active walking, demonstrating that PD-related alterations in gaze control translate, to some extent, across tasks of varying complexity (i.e. from simple seated to adaptive gait) (Gibbs et al., 2024; Tsitsi et al., 2023; Yu et al., 2016). If this observation is robust, fixation duration and saccade amplitude observed in daily life could serve as markers of visuomotor dysfunction in PD (Gibbs et al., 2024). The ability to measure such outcomes using emerging wearable eye-tracking technology (e.g. augmented reality glasses) raises the possibility of quantifying visuomotor function and generating meaningful outcomes that could be used to track changes over time.

### Associations between affect and oculomotor control

Our exploratory analyses indicated that higher gait-specific arousal was associated with reduced saccade amplitude, while greater depressive symptoms were associated with longer fixation durations. These relationships were not observed for general symptoms of anxiety.

Increased arousal has been shown to influence neural thresholds for saccade initiation and amplitude scaling via modulation of brainstem and basal ganglia circuits (Pelzer et al., 2020; Pretegiani & Optican, 2017). It is also possible that increased arousal while walking is related to heightened sensitivity to postural threat and associated adoption of visuomotor control strategies that serve to minimise potentially destabilising saccades. Similarly, depressive symptoms are associated with slower processing speed and reduced cognitive flexibility, which may manifest as reduced visual exploration and longer fixation durations (Armstrong & Olatunji, 2012; Buyukdura et al., 2011). Although the relationships observed here should be interpreted cautiously, they highlight the potential interaction between affective states, mood disorders and visuomotor control in PD.

### Implications

The present findings suggest that reduced visual exploration during walking in pwPD is not clearly related to FOG pathology or global motor impairment, but rather to functional balance capacity. This distinction may help reconcile the inconsistent picture that has emerged from previous work comparing visual search behaviour between PD subgroups defined by FOG status (Vanegas-Arroyave et al., 2022). The current results suggest that significant differences previously observed between pwPD with and without FOG may have been driven, at least in part, by between-group differences in balance ability rather than by FOG pathology itself. Consistent with this view, case-series evidence indicates substantial within-freezer heterogeneity in fixation and blink behaviour around freezing episodes, rather than a single stereotyped gaze signature (Kondo & Muroi, 2025). These observations point to the substantial variability not only in the manifestation of PD and FOG subtypes (Ehgoetz Martens et al., 2018), but also in the cognitive and affective factors, including attentional control, anxiety, and fear of falling, that are known to influence eye movements and visual search during locomotion (Young & Mark Williams, 2015). Taken together, these considerations highlight the need for more detailed phenotypic profiling to identify the factors driving maladaptive changes in eye movements and associated movement planning, and thereby to identify viable targets for future rehabilitation.

Outside fixation frequency may provide a behavioural index of visuomotor capacity during walking, reflecting the extent to which individuals are able to disengage visual attention from immediate stepping constraints (Ellmers et al., 2016; Russo et al., 2025). Because this measure does not depend on specific environmental features, it may offer a more generalisable indicator of functional locomotor stability than task-specific proactive gaze measures.

From a clinical perspective, assessment of eye movement behaviour during walking may provide complementary information to traditional gait and balance measures. Interventions targeting balance function or confidence may indirectly influence visual exploration patterns, potentially supporting more adaptive visuomotor strategies during locomotion. Preliminary evidence suggests that physiotherapy incorporating gaze training can improve balance and functional mobility in pwPD, supporting the potential relevance of visuomotor mechanisms in rehabilitation (Mildner et al., 2024).

### Limitations

Several limitations should be considered. First, pwPD were assessed “ON” medication, which is likely to have reduced the magnitude of motor and visuomotor impairments (Curtze et al., 2015). Second, the experimental task involved a relatively predictable spatial structure, which may have reduced the requirement for extended proactive visual search. More complex or unpredictable environments (e.g. changing the position and/or orientation of stepping targets) may be required to more comprehensively characterise anticipatory gaze behaviour in PD. Finally, the cross-sectional design limits inference regarding causal relationships between balance ability and visual search behaviour.Longitudinal studies are required to determine whether narrowing of visual exploration emerges as balance declines, or whether both reflect shared underlying disease mechanisms.

## 5. Conclusion

People with Parkinson’s disease demonstrated altered visual exploration during adaptive walking, characterised by longer fixation durations, reduced saccade amplitudes, and fewer fixations toward task-irrelevant outside areas. Fewer outside fixations were associated with poorer balance ability but were not related to FOG severity or overall motor impairment. These findings indicate that narrowing of visual exploration during walking is linked more closely to functional balance constraints than to freezing pathology. Moreover, outside fixation frequency may reflect spare visuomotor capacity during locomotion and may provide a relevant behavioural marker of functional stability in PD. Assessing oculomotor behaviour during walking may therefore offer insight into how balance control, sensorimotor processing, and affective factors interact to shape locomotor strategies in everyday environments. Future work should examine how visual exploration changes across disease progression and determine whether interventions targeting balance or gait-specific anxiety can modify visual search behaviour and improve functional mobility in PD.

## Data Availability

All data produced in the present study are available upon reasonable request to the authors

## List of abbreviations

AOI: Area of interest
CMP: Conscious movement processing
FDR: False discovery rate (Benjamini–Hochberg)
FOG: Freezing of gait
G-SAP: Gait-Specific Attentional Profile
HADS: Hospital Anxiety and Depression Scale
HADS-A: Hospital Anxiety and Depression Scale – Anxiety subscale
HADS-D: Hospital Anxiety and Depression Scale – Depression subscale
HC: Age-matched healthy controls
I-VT: Velocity-Threshold Identification (gaze event classification)
MDS-UPDRS-III: Movement Disorder Society – Unified Parkinson’s Disease Rating Scale, Part III (Motor Examination)
Mini-BESTest: Mini Balance Evaluation Systems Test
MoCA: Montreal Cognitive Assessment
NFOG-Q: New Freezing of Gait Questionnaire
PI: Processing inefficiency
pwPD: People with Parkinson’s disease
TI: Task-irrelevant worry
TT: Turn target

## Acknowledgments

The authors would like to thank the participants, the people who accompanied them in the laboratory, and the members of Parkinson’s UK branches.

