## Supplementary Tables for "Exploring characteristics of visual search in older adults and people with Parkinson’s during adaptive gait"

### Supplementary Materials

*Table 1: Group comparisons for eye movements variables of healthy controls (HC) and people with Parkinson's disease (pwPD) group. * p < 0.05, ** p < 0.01, *** p < 0.001.*

| Variable | HC (Mean±SD) | pwPD (Mean±SD) | p value | Effect size |
|---|---|---|---|---|
| Percentage of total fixations duration (outer obstacle) | 0.59 ± 0.75 | 1.49 ± 2.46 | 0.4971 | -0.104 |
| Percentage of total fixations duration (inner obstacle) | 0.85 ± 1.0 | 1.11 ± 2.04 | 0.8811 | 0.024 |
| Percentage of total fixations duration (TT) | 8.18 ± 6.37 | 8.89 ± 6.44 | 0.5597 | -0.09 |
| Percentage of total fixations duration (platform) | 65.86 ± 30.07 | 75.53 ± 27.15 | 0.1124 | -0.243 |
| Percentage of total fixations duration (outside) | 34.14 ± 30.07 | 24.47 ± 27.15 | 0.1124 | 0.243 |
| Average duration of fixations | 291.99 ± 70.44 | 391.47 ± 125.59 | **0.0002*** ** | -0.925 |
| Average amplitude of saccades | 9.75 ± 2.39 | 8.19 ± 2.7 | **0.0249*** | 0.603 |
| Number of fixations platform (per second) | 1.56 ± 0.71 | 1.43 ± 0.57 | 0.4284 | 0.209 |
| Number of fixations outside (per second) | 0.82 ± 0.64 | 0.49 ± 0.54 | **0.0234*** | 0.347 |

*Table 2: The correlation results between eye movement variables and Mini-Best, NFOGQ (with and without controlling for MDS-UPDRS-III).*
*Note: The Benjamini-Hochberg FDR method adjusts each p-value by multiplying it by (m/i), where m = total number of tests and i = rank. However, the procedure also enforces monotonicity - adjusted p-values cannot decrease as you move down the ranked list. When a calculated adjustment would be smaller than the previous one, it gets set equal to the previous value instead. This creates identical adjusted p-values.*

| | | Without Controlling for MDS-UPDRS-III | | | With Controlling for MDS-UPDRS-III | | |
|---|---|---|---|---|---|---|---|
| **Variable** | **Questionnaire** | **N** | **r** | **p-value (FDR)** | **N** | **Partial r** | **p-value (FDR)** |
| Percentage of total fixations duration (outer obstacles) | Mini-Bestest | 32 | -0.082 | 0.655 | 32 | 0.060 | 0.750 |
| Percentage of total fixations duration (inner obstacles) | Mini-Bestest | 32 | -0.400 | 0.055 | 32 | -0.411 | 0.051 |
| Percentage of total fixations duration (TT) | Mini-Bestest | 32 | -0.265 | 0.250 | 32 | -0.257 | 0.286 |
| Percentage of total fixations duration (outside) | Mini-Bestest | 32 | 0.451 | **0.033** | 32 | 0.450 | **0.039** |
| Average duration of fixations | Mini-Bestest | 32 | 0.083 | 0.655 | 32 | 0.104 | 0.674 |
| Average amplitude of saccades | Mini-Bestest | 32 | 0.190 | 0.417 | 32 | 0.133 | 0.667 |
| Number of fixations outside (per second) | Mini-Bestest | 32 | 0.512 | **0.019** | 32 | 0.501 | **0.029** |
| Percentage of total fixations duration (outer obstacles) | NFOG-Q | 18 | 0.093 | 0.919 | 18 | 0.116 | 0.920 |

| | | | | | | | |
|---|---|---|---|---|---|---|---|
| Percentage of total fixations duration (inner obstacles) | NFOG-Q | 18 | 0.395 | 0.736 | 18 | 0.438 | 0.549 |
| Percentage of total fixations duration (TT) | NFOG-Q | 18 | -0.145 | 0.919 | 18 | -0.119 | 0.920 |
| Percentage of total fixations duration (outside) | NFOG-Q | 18 | -0.009 | 0.971 | 18 | -0.022 | 0.933 |
| Average duration of fixations | NFOG-Q | 18 | 0.251 | 0.737 | 18 | 0.283 | 0.920 |
| Average amplitude of saccades | NFOG-Q | 18 | -0.253 | 0.737 | 18 | -0.184 | 0.920 |
| Number of fixations outside (per second) | NFOG-Q | 18 | -0.068 | 0.919 | 18 | -0.060 | 0.933 |
| Percentage of total fixations duration (outer obstacles) | G-SAP CMP | 34 | 0.021 | 0.906 | 34 | -0.034 | 0.852 |
| Percentage of total fixations duration (inner obstacles) | G-SAP CMP | 34 | 0.071 | 0.816 | 34 | 0.074 | 0.795 |
| Percentage of total fixations duration (TT) | G-SAP CMP | 34 | 0.069 | 0.816 | 34 | 0.075 | 0.795 |
| Percentage of total fixations duration (outside) | G-SAP CMP | 34 | -0.194 | 0.477 | 34 | -0.185 | 0.530 |
| Average duration of fixations | G-SAP CMP | 34 | -0.287 | 0.477 | 34 | -0.306 | 0.530 |
| Average amplitude of saccades | G-SAP CMP | 34 | -0.223 | 0.477 | 34 | -0.209 | 0.530 |
| Number of fixations outside (per second) | G-SAP CMP | 34 | -0.198 | 0.477 | 34 | -0.193 | 0.530 |

*Table 3: The correlation results of average saccade amplitude and fixation duration with MOCA, G-SAP, and HADS.*
*Note: The correlation coefficients reported in this correlation table differ from those presented in Table 3 because the analyses were conducted on slightly different participant samples.*

| | Average duration of fixations | | | Average amplitude of saccades | | |
|---|---|---|---|---|---|---|
| **Questionnaire** | **N** | **r** | **p value (FDR)** | **N** | **r** | **p value (FDR)** |
| G-SAP Arousal | 35 | -0.059 | 0.738 | 35 | -0.535 | **0.003** |
| G-SAP TI | 35 | 0.147 | 0.598 | 35 | -0.349 | 0.060 |
| G-SAP CMP | 35 | -0.270 | 0.348 | 35 | -0.261 | 0.130 |
| HADS-D | 34 | 0.387 | **0.048** | 34 | -0.276 | 0.227 |
| HADS-A | 34 | -0.045 | 0.801 | 34 | -0.055 | 0.756 |

*Table 4： The correlations result between Mini-BEST scores and eye movements variables in HC group. (FDR adjusted)*

| Variable | r | CI95% | p-value adjusted |
|---|---|---|---|
| Percentage of total fixations duration outer obstacles | -0.4652 | [-0.760, -0.010] | 0.3132 |
| Percentage of total fixations duration inner obstacles | 0.2653 | [-0.210, 0.640] | 0.6356 |
| Percentage of total fixations duration TT | 0.1415 | [-0.330, 0.560] | 0.9551 |
| Number of fixations outside (per second) | 0.0169 | [-0.440, 0.470] | 0.9551 |
| Percentage of total fixations duration outside | 0.0139 | [-0.440, 0.470] | 0.9551 |
| Average duration of fixations | 0.0718 | [-0.400, 0.510] | 0.9551 |
| Average amplitude of saccades | 0.3083 | [-0.170, 0.670] | 0.6356 |

*Table 5: Group comparison results between healthy controls (HC) and people with Parkinson's disease (pwPD) in G-SAP and HADS.*

| Variable | Test | Statistic | p-value | Effect Size |
|---|---|---|---|---|
| G-SAP Arousal | Mann-Whitney U | 118.5 | **<0.001** | 0.68 |
| G-SAP TI | Mann-Whitney U | 184.5 | **0.001** | 0.50 |
| G-SAP CMP | Mann-Whitney U | 101.5 | **<0.001** | 0.72 |
| G-SAP PI | Mann-Whitney U | 212 | **0.007** | 0.42 |
| HADS-D | Mann-Whitney U | 276.5 | 0.163 | 0.23 |
| HADS-A | t-test | -2.093 | **0.042** | -0.56 |
